# Age-Sex based estimates of risk of death from COVID Infection in Adult Cancer Patients

**DOI:** 10.1101/2020.03.18.20038067

**Authors:** Matt Williams, Ella Mi, Kerlann Le Calvez, Jiarong Chen, Lillie Pakzad-Shahabi, Seema Dadhania, James Wang, Andrew L.K.Ho, Simon Rabinowicz

## Abstract

**Background:** During the coronavirus disease 2019 (COVID) pandemic, various organisations have produced management guidance for cancer patients and the delivery of cytotoxic chemotherapy, but none offer estimates of risk, or the potential impact across populations.

**Methods:** We combine data from four countries to produce pooled age-banded Case Fatality Rates (CFRs), calculate the sex-difference in survival and use data from four recent studies to convert CFRs into age-sex stratified Infection Fatality Rates (IFRs). We estimate the additional risk of death in cancer patients, and in those receiving chemotherapy. We illustrate the impact of these by considering the impact on a national incident cancer cohort and present some clinical scenarios.

**Results:** We obtained data based on 412,985 cases and 41,854 deaths. The pooled estimate for IFR was 0.92%. Age-related IFRs for patients with cancer range from 0.01% to 29%, and higher in patients receiving chemotherapy. The risk is significantly higher in men than women. 40% of all male and 32% of all female patients with a new diagnosis of cancer this year have an IFR of ≥ 5%.

**Conclusions:** Older male patients are at a higher risk of death with COVID infection. Patients with cancer are also at higher risk, as are those who have recently received chemotherapy. We provide well-founded estimates to allow patients and clinicians to better balance these risks, and illustrate the wider impact in a national incident cohort.

**FUNDING & DISCLOSURES:** MW receives funding from the Imperial/ NIHR BRC; SD receives funding from the IC/ICR CRUK Major Centre; LPS receives funding from Brain Tumour Research and the Brain Tumour Research Campaign. JC is supported by the Guangdong International Young Research Talents Training Programme for Postdoctoral Researchers. The funders had no role in study design, data collection, data analysis, data interpretation, or writing of the report. The corresponding author had full access to all the data in the study and had final responsibility for the decision to submit for publication. Code, data and appendicies are available at: https://gitlab.com/computational.oncology/covidcancerrisk

**HIGHLIGHTS:**

- We report case and infection fatality rates based on a large multi-national cohort
- We provide sex and age-specific estimates of risk
- We provide estimates of additional risk for patients with cancer to allow patients and clinicians to balance risk and benefit

## INTRODUCTION

The world is experiencing a pandemic caused by severe acute respiratory syndrome coronavirus 2 (SARS-CoV-2; COVID). Although the overall case fatality rate (CFR) is lower than some other recent respiratory infections, the widespread pattern of infection puts many more people at risk [26]. Patients with a range of co-morbidities are at increased risk of harm [9], including those with cancer who are more susceptible to infection because of their systemic immunosuppressive state, owing to the malignancy and anticancer treatments [11]. While there are now a range of guidelines and prioritisation approaches, [7, 16], none of these address the issue of estimating risks in cancer patients.

The available data indicates a clear age effect in the risk of death from COVID [21, 28], and a higher risk of death in men compared to women. Data from 72314 cases from the Chinese Center for Disease Control and Prevention report a CFR of 5.6% in patients with cancer [26]. There is one small series of 18 patients with cancer, which suggests higher risks of intubation or death, but it is too small to draw robust conclusions. One very recent case control study also suggests higher risks of death or ITU admission in patients with COVID who have a history of cancer, compared to COVID-infected non-cancer controls [5]. There are several studies indicating an increased risk of death in patients with cancer or those who are immunosuppressed who are infected during influenza pandemics [19]. The risk estimates vary considerably, and probably represent both biological and methodological variation.

Most recent work has presented outcomes as case fatality rates (defined as the number of deaths ÷ the number of cases). These are easy to calculate, but are subject to ascertainment bias (when only more ill patients are tested for the disease) and the effect of the delay between infection and death, which makes early estimates of CFR prone to under-estimating true death rates [8]. We would ideally like to know the infection fatality rate (IFR; i.e. the proportion of those infected who die) but a robust estimate requires near-universal testing of "cases" (i.e. those who are obviously unwell), those with mild symptoms, and those who are asymptomatic. This typically requires a "closed" population where we can assess both cases and infection rates. Such situations occur when patients are screened after travel home from an infected area, or when an outbreak occurs in an isolated population. Where possible, IFRs should be preferred to CFRs because they provide a better estimate of the true population risk, accepting that most of the population are likely to be infected, but not all become "cases" (i.e. not become unwell enough to come to medical attention).

In this paper we estimate age-sex based IFRs for COVID infection, and the additional risk for cancer patients. We use that to calculate age-sex specific IFRs in cancer patients, and compare those to benefits from chemotherapy in some common clinical scenarios. We assess the impact of COVID-associated mortality in a national incident cancer population across different tumour types.

## METHODS

### Estimating IFR

We identified all national data which provided case fatality rates in 10 year (or more specific) age bands, where there were national reports of ≥ 100 deaths and a reference national age distribution. For each nation, we identified which age group had the highest case ascertainment rate (i.e. highest proportion of that age band diagnosed as COVID cases). In line with previous work [21] we assumed a uniform infection rate across age groups, adjusted CFRs in other age groups assuming that lower proportions of testing represented under-ascertainment of cases, but not deaths, to generate adjusted CFRs, and then combined these, weighted by number of deaths, to produce pooled adjusted CFRs (Appendix A). We calculated sex-associated difference in risk of death due to COVID, calculating an odds ratio for risk of death due to COVID for males (relative to females) based on pooled international data [1]. Assuming a 1:1 male:female ratio among COVID cases, which is supported by worldwide data on cases, and that differential risk between sexes was independent of age, which we confirmed by calculating age-stratified ORs for sex where age-sex stratified data on cases and deaths were available, we calculated age-sex stratified estimates of CFR. We converted CFRs into IFRs by combining CFR:IFR ratios from sources which either used closed populations with near universal ascertainment rates, or attempted to close the population through statistical methods, and which provided rates of both cases and infections ascertained using swab-based PCR (rather than serological) methods to obtain final age-sex stratified estimates of IFR for death due to COVID (Appendix C).

### Estimating risks in patients with cancer

We conducted a rapid systematic review to estimate risk of harm (including death) in patients with cancer during viral outbreaks. We searched three bibliographic databases on the 13^th^ March 2020 (Appendix D). We conducted a second rapid systematic review on the 1^st^ May 2020 to identify the risk of severe harm and/or death in patients with cancer during the COVID pandemic (Appendix E) ensuring we covered the same terms as a recent review [6]. For both reviews, we supplemented with additional resources found through grey literature and web searches, including pre-prints, removed duplicates and screened titles and abstracts for relevance; those thought to be relevant had full-text extracted and reviewed by two reviewers (LPS and JC). Studies were included in the final analysis if they reported some measure of risk (normally as an Odds Ratio) from cancer or chemotherapy in the context of a viral pandemic. We extracted data on study population, methodology and measures, and conducted a random-effects meta-analysis of the Odds Ratio (OR) for death.

We used age-sex-stratified IFR and the estimate of additional risk of death with cancer alone and cancer with chemotherapy to generate expected age-sex-stratified IFRs for COVID patients who also have cancer and chemotherapy. We plotted estimated IFRs based on age-sex alone, age-sex with cancer and age-sex with cancer and chemotherapy.

### Clinical Scenarios and Risk-benefit analysis

We constructed clinical scenarios by identifying clinical decision tools or key clinical trials in the curative setting in three tumour sites. We extracted data on overall survival and absolute difference in overall survival at timepoints specified. We analysed risk-benefit of giving chemotherapy in these clinical scenarios, comparing survival benefit from chemotherapy to increased risk of death from COVID due to chemotherapy if the patients in these scenarios did acquire COVID (i.e. risk of infection of 1). Increased risk of death from COVID was calculated from excess risk of death from COVID due to chemotherapy adjusted for total risk of death from COVID with cancer and chemotherapy and overall survival from cancer with chemotherapy. We extended the risk-benefit analysis by applying it to a putative population of cancer patients of defined age, sex, cancer site and treatment intent, informed by one of these clinical scenarios. Details of derivation of the equation used and the worked population example can be found in Appendix B.

### Population-level impact

We obtained data on all new diagnoses of invasive cancer in the UK between 2016 - 2018 inclusive, and calculated average annual figures. We calculated age-sex IFRs for every tumour site and examined the numbers and proportions of patients with each cancer where the IFR exceeded 2.5, 5, 10 and 20% (Appendix F).

### Roles within the study

MW instigated, designed and oversaw the work, and wrote the initial draft of the manuscript. EM wrote code to generate IFRs and designed the model for risk benefit analysis. KLC calculated adjusted CFRs. JC and LPS designed, conducted and reported the rapid systematic reviews. SD designed, identified and extracted data for the clinical scenarios. JW designed and carried out the IFR calculations for incident populations. AH revised the IFR: CFR data and calculation, and sourced additional national data. SR contributed to manuscript review, sourcing national data and additional calculations. All authors contributed to and reviewed the final manuscript before submission.

## RESULTS

Risk of death with COVID infection increases substantially with age, and is higher in men. The risks are higher in patients with cancer and chemotherapy, and this risk is multiplicative, and so the absolute increase in risk is larger in older and male patients. Figure 1 shows the estimated IFRs for death due to COVID age-sex band, divided by contribution from age-sex, cancer and chemotherapy.

**Figure 1.**
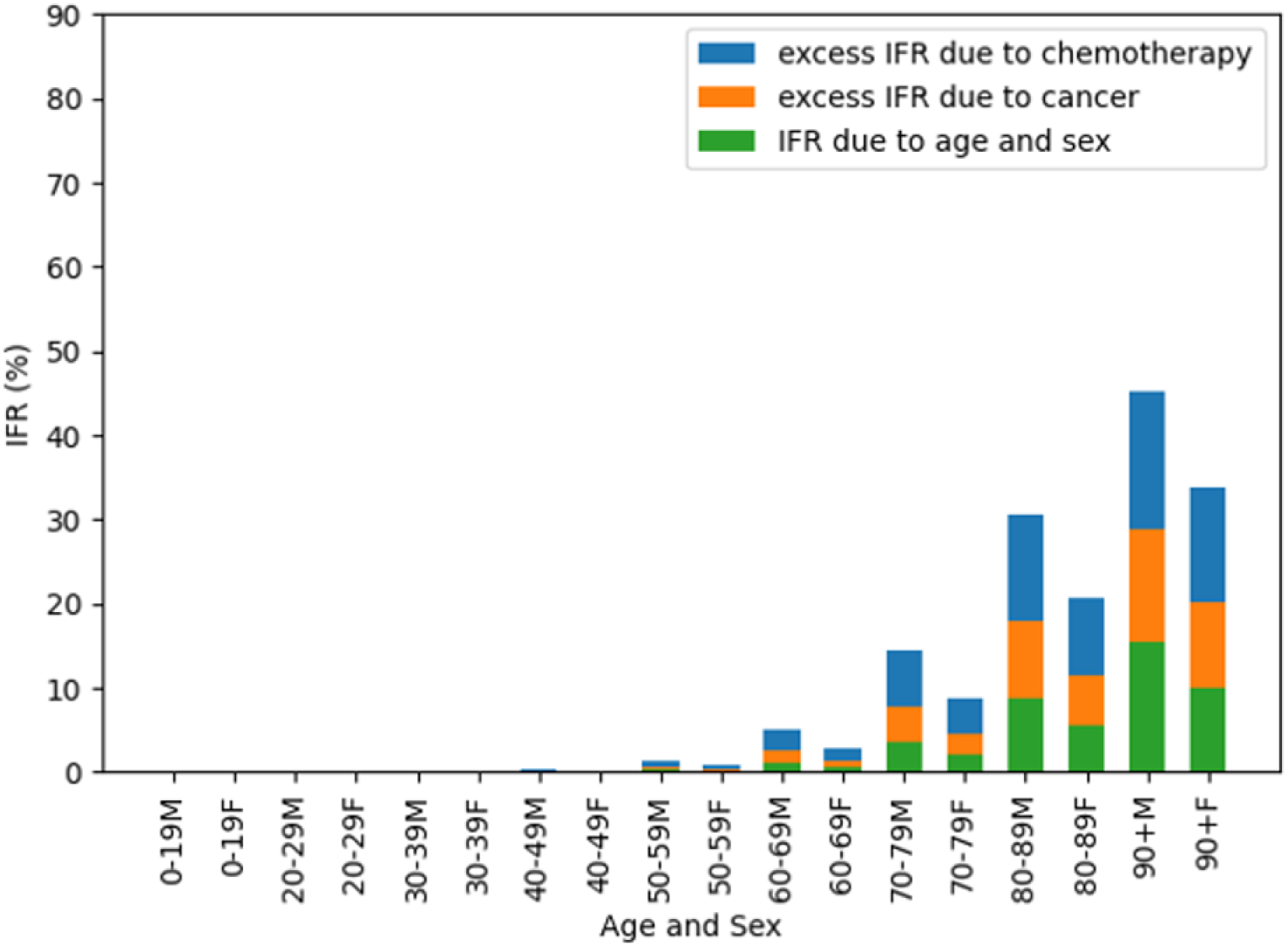
Age-sex IFRs and contribution by age-sex, cancer and chemotherapy.

We included data on 412,985 cases and 41,854 deaths from four different countries (China, Italy, the Netherlands and Spain) to generate our estimates of age-related CFR. The age-group with the highest ascertainment ratio was the 90+ year old group. Adjustment of age-stratified CFRs for under-ascertainment reduced the CFRs considerably, especially in the 50 – 70 year old age group (Table 1). The odds ratio for death for males (vs females) was 1.77 (95% CI 1.73 - 1.80), based on data on 819,145 cases and 57,814 deaths from 30 countries [1]. This OR was used to convert adjusted age-stratified CFRs to age-sex stratified CFRs (Table 2). We identified four studies that reported both CFRs and IFRs in populations with near complete case ascertainment based on modelling infection rates in travellers, travellers returning from Wuhan to Japan, infections on the Princess Diamond cruise ship, and a COVID outbreak in a Seattle care home [12, 17, 18, 21]. Based on these we calculated an IFR:CFR ratio of 0.48 (95% CI 0.47 - 0.48). This led to the calculation of age and age-sex stratified IFRs in Tables 1 and 2.

**Table 1.** Deaths, crude CFR and adjusted CFR by age band for four countries individually and pooled.

| Age | Italian deaths | Crude Italian CFR | Adjusted Italian CFR | Dutch deaths | Crude Dutch CFR | Adjusted Dutch CFR | Spanish deaths | Crude Spanish CFR | Adjusted Spanish CFR | Chinese deaths | Adjusted Chinese CFR | Combined CFR | Combined IFR |
| --- | --- | --- | --- | --- | --- | --- | --- | --- | --- | --- | --- | --- | --- |
| 0-19 | 0 | 0.0% | 0.00% | 1 | 0.21% | 0.00% | 5 | 0.36% | 0.00% | 1 | 0.02% | 0.00% | 0.00% |
| 20-29 | 0 | 0.0% | 0.00% | 3 | 0.09% | 0.01% | 25 | 0.31% | 0.03% | 7 | 0.06% | 0.03% | 0.02% |
| 30-39 | 44 | 0.3% | 0.04% | 8 | 0.24% | 0.02% | 50 | 0.35% | 0.05% | 18 | 0.15% | 0.06% | 0.03% |
| 40-49 | 209 | 0.9% | 0.13% | 19 | 0.44% | 0.04% | 138 | 0.61% | 0.10% | 38 | 0.30% | 0.13% | 0.06% |
| 50-59 | 834 | 2.6% | 0.54% | 112 | 1.57% | 0.22% | 401 | 1.44% | 0.32% | 130 | 1.25% | 0.52% | 0.25% |
| 60-69 | 2529 | 9.8% | 2.07% | 388 | 7.54% | 0.92% | 1152 | 4.87% | 1.22% | 309 | 3.99% | 1.88% | 0.90% |
| 70-79 | 6723 | 24.3% | 6.79% | 1308 | 23.40% | 4.24% | 3375 | 15.01% | 4.85% | 312 | 8.61% | 5.99% | 2.85% |
| 80-89 | 9413 | 30.8% | 16.27% | 1966 | 30.19% | 14.42% | 5502 | 24.05% | 13.34% | 208 | 13.40% | 15.08% | 7.18% |
| 90+ | 3409 | 27.7% | 27.65% | 760 | 29.60% | 29.60% | 2457 | 24.95% | 24.95% | NA | NA | 26.87% | 12.80% |
| Total | 23161 | 13.1% | 2.28% | 4565 | 11.89% | 1.30% | 13105 | 8.57% | 1.56% | 1023 | 1.38% | 1.93% | 0.92% |

**Table 2.** Age-sex CFRs and IFRs in baseline populations, in patients with cancer and in patients with cancer receiving chemotherapy.

| Age/ Sex Group | CFR by age and sex | IFR by age and sex | IFR with Cancer | IFR with Cancer + Chemotherapy |
| --- | --- | --- | --- | --- |
| 0-19 M | 0.01% | 0.00% | 0.01% | 0.01% |
| 0-19 F | 0.00% | 0.00% | 0.00% | 0.01% |
| 20-29 M | 0.04% | 0.02% | 0.05% | 0.09% |
| 20-29 F | 0.02% | 0.01% | 0.03% | 0.05% |
| 30-39 M | 0.07% | 0.03% | 0.07% | 0.15% |
| 30-39 F | 0.04% | 0.02% | 0.04% | 0.09% |
| 40-49 M | 0.17% | 0.08% | 0.18% | 0.36% |
| 40-49 F | 0.09% | 0.05% | 0.10% | 0.20% |
| 50-59 M | 0.66% | 0.32% | 0.70% | 1.41% |
| 50-59 F | 0.37% | 0.18% | 0.40% | 0.81% |
| 60-69 M | 2.39% | 1.14% | 2.51% | 4.97% |
| 60-69 F | 1.37% | 0.65% | 1.44% | 2.89% |
| 70-79 M | 7.57% | 3.60% | 7.69% | 14.51% |
| 70-79 F | 4.42% | 2.10% | 4.57% | 8.89% |
| 80-89 M | 18.67% | 8.89% | 17.87% | 30.70% |
| 80-89 F | 11.48% | 5.47% | 11.43% | 20.80% |
| 90+ M | 32.42% | 15.44% | 28.93% | 45.32% |
| 90+ F | 21.32% | 10.15% | 20.13% | 33.91% |
| Average Male | 2.45% | 1.17% | 2.57% | 5.09% |
| Average Female | 1.40% | 0.67% | 1.47% | 2.96% |

We identified 850 distinct articles for the first rapid review; we used data on 1318 patients from 4 studies based on patients during influenza pandemics to calculate a pooled OR for death of 3.7 (95% CI 1.5 - 9.1) [4, 10, 13, 20] (Appendix D). The second, COVID-specific rapid review identified 418 distinct articles from 3 search engines and an additional 1 through a web search; we identified 1 case-control study that reported odds ratios for the risk of death in cancer patients infected with COVID (Appendix E). This study reported elevated risk of death and ITU admission in 105 cancer patients compared with 536 matched controls, with an OR for death in cancer patients of 2.34 (95% CI 1.15 - 4.77), OR of death in cancer without active treatment of 2.23 (0.98 - 5.11) and OR of death in cancer with chemotherapy of 4.54 (1.21 - 16.99) [5]. We used these latter two ORs to convert baseline age-sex stratified IFRs to age-sex stratified IFRs in cancer +/- chemotherapy in Table 2.

Table 3 shows three clinical scenarios in which chemotherapy is given in the curative setting to patients (of specified age and sex) with breast, lung and brain tumours. In general, survival benefits for cancer due to chemotherapy were 5-10% at 5-10 years, compared to increased risk of death from COVID due to chemotherapy ranging from 0-3% if the patients acquired COVID. Extending this to a population, Figure 2 presents expected death and survival among 100 men aged 60-69 with lung cancer treated with adjuvant chemotherapy to further illustrate the trade-off between survival benefit from cancer and increased COVID mortality conferred by chemotherapy.

**Table 3.** Clinical scenarios to illustrate balance of survival benefit and increased risk of death from Covid with chemotherapy.

| Tumour Group | Clinical Decision Tool/Evidence | Case History | Treatments and benefit | Risk Benefit Analysis |
| --- | --- | --- | --- | --- |
| Breast | NHS PREDICT (breast cancer) | 71-year-old post-menopausal woman with a symptomatic pT3 (60 mm) N0 M0 ER+ HER2- G3 IDC who has a WLE + SLNB. | Survival at 10 years:<br>Surgery only = 45%<br>+ hormone therapy = 54%<br>+ chemotherapy = 54%<br>+ bisphosphonates = 63% | There is a 4.8% benefit at 10 years with adjuvant FEC (second generation) chemotherapy.<br>This compares the with increased risk of death if she acquires COVID-19 of 2.4% |
| Lung | LACE Meta-analysis | 68-year-old male with resected Stage IIIA NSCLC, EGFR wildtype; PDL1 <1% | Survival at 5 years:<br>Surgery only = 43.4%<br>+ chemotherapy = 48.8% | There is a 5.4% survival benefit at 5 years with cisplatin-based chemotherapy.<br>This compares with the increased risk of death if he acquires COVID-19 of 1.2% |
| Brain | Stupp 2009 | 63-year-old male with WHO Grade IV resected, MGMT methylated glioblastoma multiforme | Survival at 5 years:<br>Surgery and radiotherapy = 0%<br>+ chemotherapy = 11% | There is an 11% survival benefit at 5 years with concurrent and adjuvant temozolamide<br>This compares with the increased risk of death if he acquires COVID-19 of 0.3% |

**Table 4.** Age-sex stratified incident cancer population for four common cancers, with percentage patients who exceed different IFR thresholds.

| TUMOUR SITE | Annual Cancer Population by Age and Sex with Heat Map of IFR |  |  |  |  |  |  |  |  |  |  | % Patients at IFR threshold |  |  |  |
| --- | --- | --- | --- | --- | --- | --- | --- | --- | --- | --- | --- | --- | --- | --- | --- |
|  | Heat Map: |  |  |  |  |  |  |  |  |  | Total |  |  |  |  |
| | Sex | 0-19 | 20-29 | 30-39 | 40-49 | 50-59 | 60-69 | 70-79 | 80-89 | 90+ | | $\geq 2.5\%$ | $\geq 5\%$ | $\geq 10\%$ | $\geq 20\%$ |
| BREAST | F | 0 | 89 | 691 | 2541 | 4131 | 4601 | 3381 | 2235 | 600 | 18269 | 34.0% | 15.5% | 15.5% | 3.3% |
|  | M | 0 | 1 | 2 | 6 | 18 | 32 | 37 | 25 | 4 | 124 | 79.2% | 53.1% | 23.2% | 3.2% |
| COLORECTAL | M | 14 | 36 | 123 | 254 | 701 | 1280 | 1797 | 1664 | 398 | 6266 | 82.0% | 61.6% | 32.9% | 6.3% |
|  | F | 9 | 30 | 120 | 270 | 924 | 2013 | 2490 | 1728 | 255 | 7838 | 57.1% | 25.3% | 25.3% | 3.2% |
| LUNG | M | 2 | 6 | 23 | 162 | 730 | 2163 | 3110 | 1876 | 296 | 8368 | 89.0% | 63.1% | 26.0% | 3.5% |
|  | F | 2 | 7 | 24 | 155 | 732 | 1954 | 2665 | 1703 | 337 | 7579 | 62.1% | 26.9% | 26.9% | 4.4% |
| PROSTATE | M | 1 | 0 | 3 | 174 | 1760 | 5322 | 5995 | 2510 | 397 | 16162 | 88.0% | 55.1% | 18.0% | 2.5% |
| ALL CANCERS | M | 388 | 581 | 1571 | 4514 | 10392 | 18626 | 20310 | 12035 | 2291 | 70706 | 60.2% | 40.3% | 16.3% | 2.4% |
|  | F | 320 | 508 | 946 | 1842 | 4581 | 8409 | 10114 | 6987 | 1412 | 35120 | 70.2% | 31.9% | 31.9% | 5.7% |

**Figure 2.**
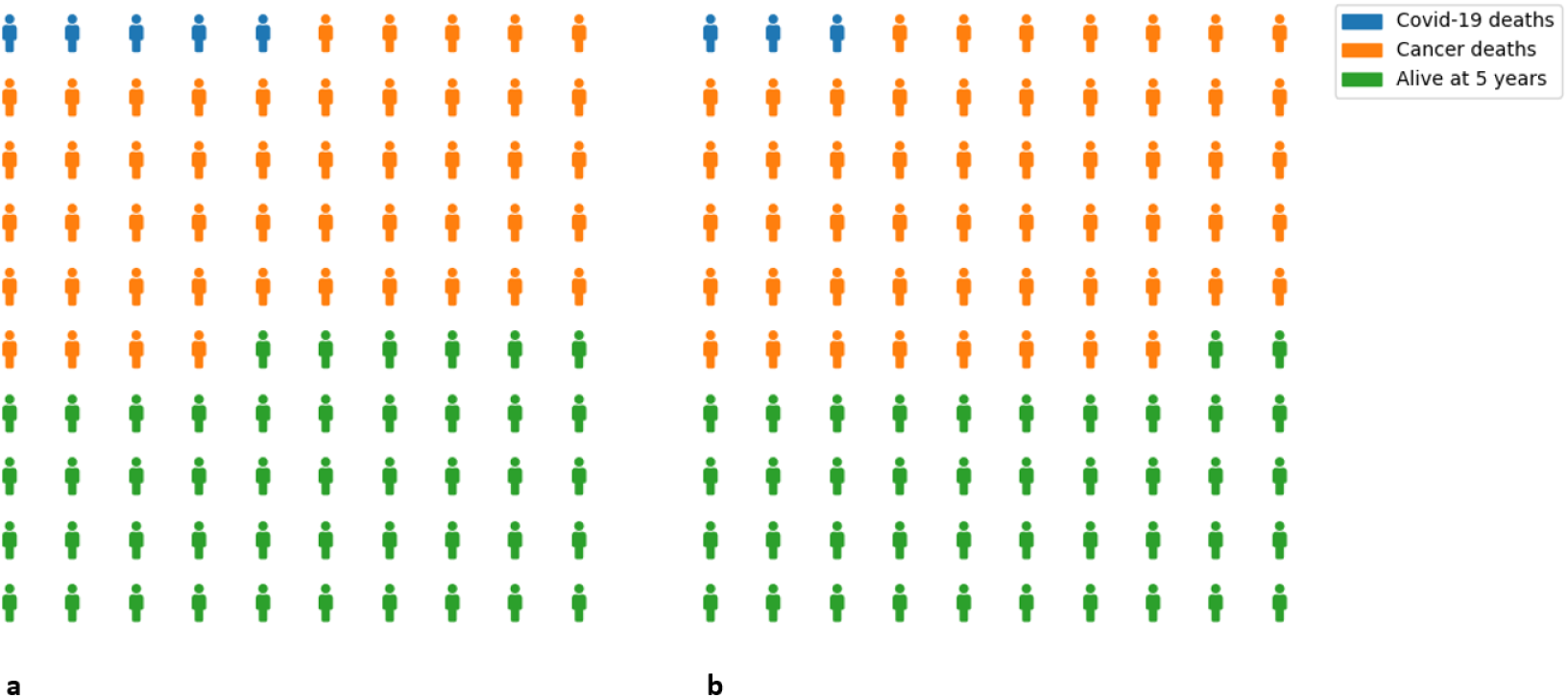
Projected deaths and survivals in 100 males aged 60-69 with lung cancer who are (a) given and (b) not given adjuvant chemotherapy, who all contract Covid-19.

## DISCUSSION

There is now clear evidence that age and sex are significant risk factors for death in patients infected with COVID. However, most of the published data is based on CFRs. Our work presents the first estimates of IFR based on pooled international data, and uses those to estimate the IFR in patients with cancer, and the additional risk when receiving chemotherapy. We use that to estimate the IFR in a national incident cancer cohort, and show that a significant minority of newly diagnosed cancer patients have a risk of COVID related death ≥ 5%. Although there are now many COVID-related cancer guidelines, this is the first that provides estimates of risk based on simple patient characteristics.

Previous work has largely presented age-based CFRs; one work has calculated age-based IFRs, based on just over 1000 deaths in patients from China. In contrast, we draw on data from > 40,000 deaths in > 400,000 cases from four countries, address the sex-difference in risk, and use a pooled estimate of the additional risk of death in patients with cancer. Our approach of adjusting CFRs to account for under-ascertainment of mild cases is similar to that used in previous work, and the conversion from CFR to IFR is simple but supported by estimates from multiple sources. Accurately estimating IFR is challenging, and is biased by under-testing of mild cases, time delay between infection, clinical symptoms and death, and under-ascertainment of deaths in the community. Our current model attempts to adjust for this, but the eventual solution relies on widespread testing of both symptomatic and asymptomatic individuals. Nonetheless, the estimates of age-sex IFR are very robust, with very small confidence intervals. Risk of death with cancer is unlikely to be already accounted for by age-sex-stratified IFRs as the prevalence of cancer among all COVID cases and deaths is negligible (1-2%).

Although we used odds ratios for increased risk of death in cancer patients from the only COVID-specifi study, the direction and magnitude of effect consistent with other work. Previous studies during viral pandemics report an increased risk of death for cancer patients [4, 10, 13, 20], and a very recent large UK COVID-specific study also shows an increased risk of death in cancer patients [25]. The estimate used in this work lies within the range of other estimates, and there are now therefore multiple sources of evidence that support the fact that patients with cancer are at higher risk of death from COVID. Although the exact figures depend on accurate parameter estimation, the clinical implications are less sensitive. The IFR due to age-sex alone in patients aged ≥ 70 is such that any of the estimates of elevated risk in cancer patients, whether COVID-specific or not, suggest that the IFR in cancer patients aged over 70 are likely to be ≥ 4% for women and 7% for men. For many patients, this is a significant increase in accepted risk levels; as a comparision, in a large national adjuvant chemotherapy cohort the 30-day mortality was 1% [22].

Other COVID-specific data also supports our conclusions. In one study of 138 patients, 40% of those with a history of cancer required ITU admission, compared to a baseline risk of 26% [23], but this included only 10 patients with cancer. A review of 18 cancer patients who developed COVID infection showed a higher risk of harm (defined as requiring ventilation in ITU or death) in 7/18 (39%) than the general population 124/1572 (8%). 12 of these 18 were being followed up after surgery, while the 4 who had undergone chemotherapy or surgery within the last month had higher risk still [14]. However, these studies do not report how many patients in each age group had each co-morbidity, and do not allow us to generate measures of risk (such as odds ratios) [3]. The exact reasons for this higher risk are unclear. Patients with cancer might be at higher risk of infection [27], and there are well described immunosupressive effects of cancer, and chemotherapy; the relative impact of each as yet is difficult to distinguish.

Although more COVID-specific data will emerge, this will take time: of data on 8250 patients who have been admitted to intensive care in the UK, only 1.9% have either metastatic cancer or haematological malignancies [2]. The systematic reviews we have performed highlight the lack of information. In particular, there is a need to distinguish between the risk due to cancer, the risk in different groups of cancer patients and the risks associated with treatment. This will require detailed, large-scale prospective data collection, and there are a variety of projects aiming to deliver this.

Our model provides age-sex stratified IFRs, based on a large pooled multi-national data from a variety of sources. We only require users to be able to specify overall survival with treatment, absolute improvement in overall survival with treatment and the patients age and sex in order to balance risk against benefit. For most younger patients, especially women, the benefits continue to outweigh the risks. However, considering the entire incident population, there is a significant risk of COVID-related death especially in older patients. This risk depends on infection, and emphasises the importance of reducing the risk of infection in newly referred patients. Given that newly diagnosed cancer patients often require significant investigations to establish a diagnosis, it is important to rationalise these and weigh them against patient-specific IFRs.

Informed consent relies on understanding the balance between risk and benefit; given that the risk associated with treatment has changed, we would suggest reconfirming informed consent in patients who started chemotherapy before the COVID pandemic and are continuing. Palliative chemotherapy may be given primarily for symptom relief, rather than to improve survival, and there are some situations where palliative radiotherapy might be substituted for chemotherapy. Radiotherapy and non-cytotoxic agents (e.g. immunotherapy and hormones) almost certainly have different risk profiles, but there are common concerns given the nosocomial pattern of disease transmission for COVID [24].

There may be additional factors to consider, such as additional co-morbidities, ethnicity, or differences in risk between different primary tumour groups. There is data to believe these may be important, but at present, there is not enough data to model these. However, our approach is flexible enough to incorporate these as they become available.

Although most of Western Europe has now passed the first peak of infections, the majority of the population remain uninfected, with estimates of only 3% of populations having been infected so far [15]. There is therefore a considerable ongoing risk which is likely to persist for many months. In contrast to outbreaks of seasonal infections, the majority of the population is expected to be infected with COVID over a short-time period (3 - 6 months), there is no pre-existing immunity or vaccine, and the case-fatality rate is approximately 5 fold higher. For those reasons, decision-making in seasonal viral outbreaks does not directly transfer to the COVID pandemic. In addition, although we have focused on death as an outcome, there are increasing reports of morbidity in survivors. Decisions the investigation and treatment of cancer patients in the context of a COVID pandemic need to be made carefully, and in light of the available data.

## Data Availability

Readers can explore data and appendicies at https://gitlab.com/computational.oncology/covidcancerrisk

## Appendices

All appendices are available online at our gitlab repository: https://gitlab.com/computational.oncology/covidcancerrisk

## Acknowledgments

Our thanks to Katie Spencer and Alice Dewdney for highlighting errors in calculations in an earlier version; to Alison Falconer for discussions about palliative chemotherapy; to those who commented on an earlier draft, and to all of those who have provided data on which this work is based.

